# Motor cortex beta oscillations reflect motor skill learning ability after stroke

**DOI:** 10.1101/2020.01.15.20017665

**Authors:** Svenja Espenhahn, Holly E Rossiter, Bernadette CM van Wijk, Nell Redman, Jane M Rondina, Joern Diedrichsen, Nick S Ward

## Abstract

Recovery of skilled movement after stroke is assumed to depend on motor learning. However, the capacity for motor learning and factors that influence motor learning after stroke have received little attention. In this study we firstly compared motor skill acquisition and retention between well-recovered stroke patients and age- and performance-matched healthy controls. We then tested whether beta oscillations (15–30Hz) from sensorimotor cortices contribute to predicting training-related motor performance.

Eighteen well-recovered chronic stroke survivors (mean age 64±8 years, range 50–74 years) and twenty age- and sex-matched healthy controls were trained on a continuous tracking task and subsequently retested after initial training (45–60 min and 24 hours later). Scalp EEG was recorded during the performance of a simple motor task before each training and retest session. Stroke patients demonstrated capacity for motor skill learning, but it was diminished compared to age- and performance-matched healthy controls. Further, although the properties of beta oscillations prior to training were comparable between stroke patients and healthy controls, stroke patients did show less change in beta measures with motor learning. Lastly, although beta oscillations did not help to predict motor performance immediately after training, contralateral (ipsilesional) sensorimotor cortex post-movement beta rebound (PMBR) measured after training helped predict future motor performance, 24 hours after training. This finding suggests that neurophysiological measures such as beta oscillations can help predict response to motor training in chronic stroke patients and may offer novel targets for therapeutic interventions.

## 1 Introduction

Stroke is a leading cause of adult disability, with lasting motor impairment being a common post-stroke outcome (Feigin *et al*., 2014). Recovery from motor impairment relies on various forms of rehabilitative training to (re)learn new or lost motor skills through repetitive practice (Krakauer, 2006; Ward *et al*., 2019). Whilst there is currently no evidence that stroke survivors lose their capacity for motor skill acquisition (Hardwick *et al*., 2017), there are considerable inter-individual differences in response to rehabilitative training, making predictions about recovery challenging (Stinear, 2010). The reasons for this clinical phenomenon are unclear. A better understanding of the underlying neurophysiological processes could therefore provide novel and important targets for improving post-stroke upper limb recovery.

The potential for plasticity in the post-stroke brain is important as it could facilitate or hinder recovery of function. Beyond the hyperacute stroke period, alterations in cortical inhibitory and excitatory mechanisms are important determinants of the potential for plasticity (Cramer, 2008, Murphy and Corbett, 2009*b*; Carmichael, 2012; Zeiler *et al*., 2013). Early stroke-induced hyperexcitability triggered by reduced GABAergic inhibition and increased glutamatergic excitation (Que *et al*., 1999) facilitates long-term potentiation (LTP) (Hagemann *et al*., 1998), downstream changes in neuronal structure (Chen *et al*., 2011), and remapping of sensorimotor functions to intact cortical areas (Takatsuru *et al*., 2009). In humans, corroborative evidence that a decrease in GABAergic inhibitory signalling after stroke is one of the key modulators of plasticity has also been obtained (Swayne *et al*., 2008; Kim *et al*., 2014; Blicher *et al*., 2015). Consequently understanding how to take advantage of post-stroke alterations in cortical inhibition and excitation to promote recovery is an important clinical and scientific goal.

Bridging the gap between cellular and behavioural accounts of post-stroke recovery, requires an appropriate biomarker reflecting underlying biological processes that predict recovery and treatment response in a way that behaviour alone cannot (Ward, 2017). Since neuronal oscillations in the beta frequency range (15-30 Hz) are fundamental for motor control (Engel and Fries, 2010) and have recently been linked to GABAergic activity in humans (Jensen *et al*., 2005; Hall *et al*., 2010, 2011; Muthukumaraswamy *et al*., 2013), properties of beta activity may provide insight into the dynamics of disease, potentially providing a clinically relevant biomarker of net inhibitory and excitatory mechanisms in human cortex. Recent evidence suggests that beta power in the sensorimotor cortex is altered after stroke, with beta activity closely tied to the degree of motor impairment (Laaksonen *et al*., 2012, Rossiter *et al*., 2014*a*; Shiner *et al*., 2015; Thibaut *et al*., 2017). Although relevant for motor control and sensorimotor pathology, and allegedly instrumental to motor learning (Boonstra *et al*., 2007; Houweling *et al*., 2008; Pollok *et al*., 2014; Espenhahn *et al*., 2019), little is known about the relationship between beta oscillations and motor learning after stroke.

Here, we explored the neurophysiological mechanisms associated with short-term motor learning after stroke in well-recovered patients. We purposefully studied well-recovered chronic stroke patients in order to assess motor learning ability independent of potentially obscuring influences of motor impairments. Since only few studies have explored post-stroke motor learning, we further investigated whether stroke patients demonstrate altered learning capability compared to healthy adults, and whether abnormal beta oscillatory activity as reported in previous studies (Rossiter *et al*., 2014*a*; Shiner *et al*., 2015) persist in patients with a low level of impairment.

## 2 Materials and methods

### 2.1 Patients and controls

Eighteen chronic stroke patients (mean age 64±8 years, range 50–74 years; for more details see **Supplementary Table 1**) with a first-time ischaemic stroke took part in the present study over two consecutive days. Two patients had to be excluded because of technical problems during data acquisition. All patients (N=16) fulfilled the following inclusion criteria: (a) suffered a stroke more than 6 months ago (chronic stage; mean time since stroke 90±50 months); (b) active range of motion around the affected wrist greater than 60° in total; (c) no reported history of other neurological or psychiatric disease; (d) no language or cognitive deficits sufficient to impair cooperation in the experiment; (e) no use of drugs affecting the central nervous system or self-reported abuse of any drugs; and (f) normal or corrected-to-normal vision. Stroke-related impairment, cognitive functioning, post-stroke fatigue and sleep were evaluated using standardized measures (see supplementary materials). As a control group, twenty age- and sex-matched healthy subjects (mean age 68±5 years, range 53–77 years) were included. Results from this healthy cohort have been published separately (Espenhahn *et al*., 2019), and here we used the exact same tasks and experimental design to investigate motor learning and beta oscillations in stroke patients. All subjects were tested between 9am and 2pm and were instructed to abstain from alcohol and caffeine for 12 hours prior to testing. The study was approved by the National Hospital for Neurology and Neurosurgery, UCL Hospitals NHS Foundation Trust and the local research ethics committee at University College London where the study was conducted. All subjects gave written informed consent in accordance with the Declaration of Helsinki.

### 2.2 Experimental design

The experimental design is illustrated in **Figure 1**A. All subjects trained with the wrist of their affected (contralesional; stroke patients) or non-dominant (controls) arm on a continuous tracking task over a single training session (40 blocks) with the aim of improving motor performance beyond pre-training levels. Motor performance was defined as the accuracy with which subjects’ wrist movement tracked the target movement (**Figure 1**B). Subjects’ motor performance was retested at two different time points: 45–60 min (retest1 on day 1; 5 blocks) and 24 hours (retest2 on day 2; 10 blocks) after initial training.

**Figure 1:**
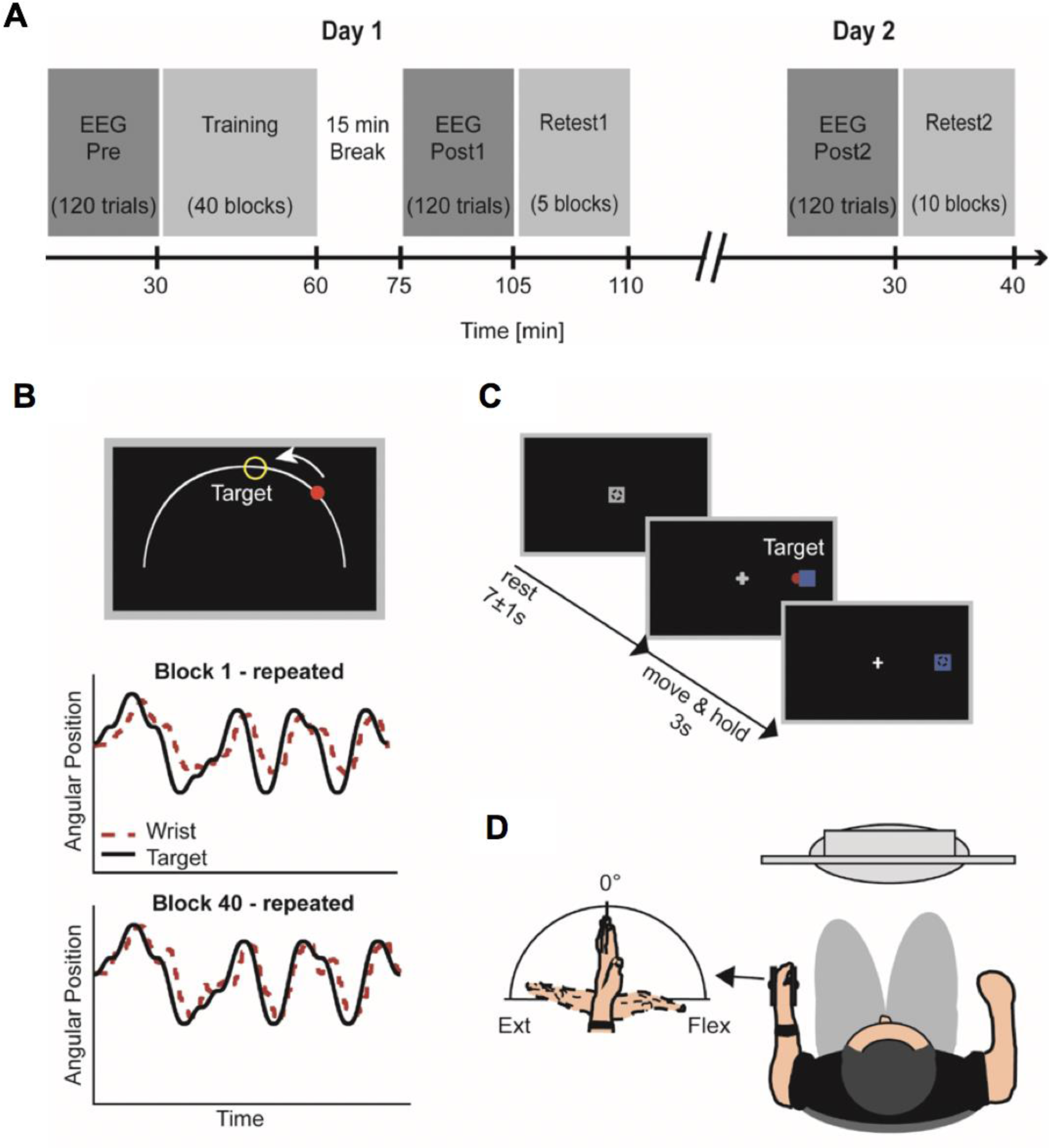
Experimental design and tasks. **A**, EEG was recorded during the performance of a simple wrist flexion / extension task (C) before (Pre) and at two time points after the training phase (Post1, Post2). Performance on the motor learning task (B) was retested after a time delay on the same day (retest1 on day 1, 45–60 min after initial training) and the following day (retest2 on day 2, 24-hours after initial training). **B**, Subjects were trained to track a target (yellow circle) moving back and forth along a fixed arc as accurately and smoothly as possible. Online visual feedback in terms of a colour change of the wrist cursor (red to green) was provided at times when the wrist cursor was located inside the circular target. Original recordings during the continuous tracking task at the beginning and end of the initial training are shown for the repeated sequence of an example patient (B, lower panel). The solid black line represents the motion of the target, while the dashed red line represents the motion of the wrist. **B**, For the simple wrist flexion / extension task, subjects were instructed to perform wrist flexion and extension to move the wrist cursor (red circle) from the initial start position (grey square) to one of two target positions (blue square) upon target presentation. The task comprised 120 trials. **D**, During both tasks, subjects sat in front of a computer monitor with their affected (patients) or non-dominant (controls) hand rested in a wrist rig that restricted movement to flexion and extension around the wrist joint. Figure adapted from (Espenhahn et al., 2019).

Electroencephalography (EEG) recorded during the performance of a simple wrist flexion / extension task (**Figure 1**C) was used to assess changes in pre-movement (resting) and movement-related beta activity before (Pre), 15 minutes after (Post1) and 24-hours after (Post2) the initial training phase.

### 2.3 Apparatus and tasks

All tasks were performed using an instrumented wrist rig (modified from (Turk *et al*., 2008)), which has been described in detail in (Espenhahn *et al*., 2019). The wrist’s angular position was continuously displayed on a computer monitor as a red circle – hereafter referred to as wrist cursor. The mid-point and maxima of a subject’s maximum active range of movement (AROM) around the wrist joint was measured and subsequently used as, respectively, start and target positions in the continuous tracking task and simple motor task. Stimuli were presented using custom software routines written in Matlab (version R2013b; The MathWorks, Inc., Natick, MA, USA).

### 2.4 Continuous tracking task

For a detailed description of the continuous tracking task, please refer to (Espenhahn *et al*., 2019). Briefly, patients were required to continuously track a circular target (in yellow) that moved back and forth along of a fixed arc through a predefined sequence of 12 positions (**Figure 1**B). Two types of sequences were randomly presented in each block, with a 3s stationary target between both; a random sequence which was only encountered once and a repeated sequence which was identical throughout training (40 blocks) and retest sessions (5 and 10 blocks). The same set of 57 difficulty-matched sequences was used across participants. Subjects were instructed to move their wrist so as to shift the red wrist cursor to match the movement of the target as ‘accurately and smoothly as possible’. Improvement on the random sequence is a measure of general skill learning, whilst any additional improvement on the repeated sequence reflects sequence-specific motor learning of the precise sequence pattern (Wulf and Schmidt, 1997). In order to ensure that the task was of equal difficulty for patients and controls at the beginning of the training and left enough room for improvement in performance, the average velocity with which the target moved along the arc was individually determined prior to training (see supplementary materials). Online visual feedback was provided during training and retest sessions and subjects received explicit verbal information about the presence of a repeated sequence along with a random sequence. However, they were not shown the repeated sequence and the target and wrist cursor trajectories did not leave a residual trail on the screen. Hence, subjects could not visualize the entire target sequence.

### 2.5 Simple wrist flexion and extension task

For a detailed description of the simple wrist flexion / extension task, please refer to (Espenhahn *et al*., 2016). Briefly, subjects performed visually cued wrist flexion and extension movements during EEG recording (**Figure 2**B). The cue to perform wrist movements was the appearance of a target at the subject’s maximum wrist flexion or extension position in a random order. Subjects were instructed to move their wrist upon presentation of the target so as to shift the red wrist cursor from the central start position to match the position of the target in a ‘quick and discrete’ movement. The target position was displayed for 3s. Once subjects returned to the initial start position, the next cue was delivered following a delay of 7±1s. The task comprised 120 trials.

**Figure 2:**
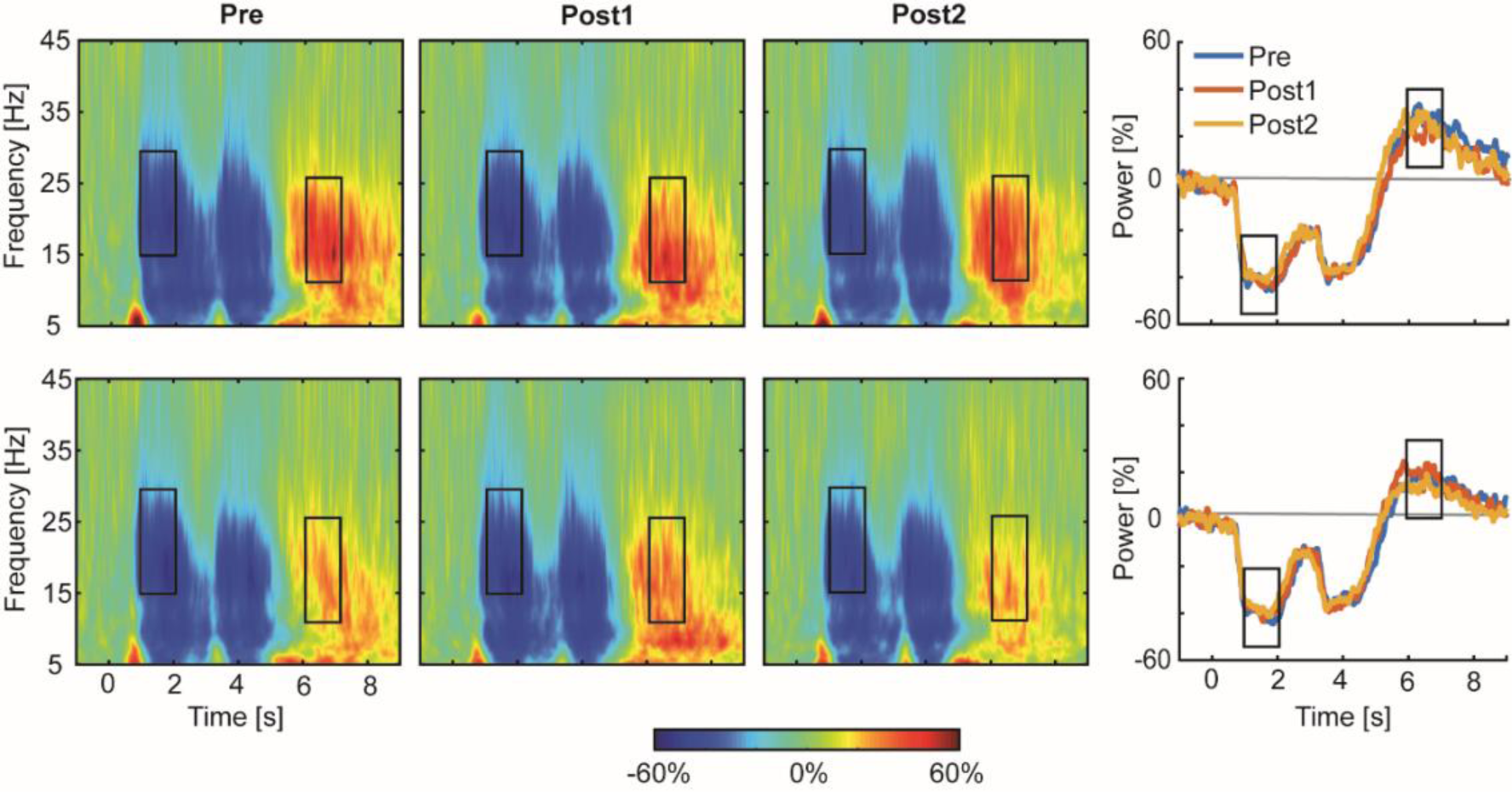
Movement-related changes in spectral power in chronic stroke patients. Time-frequency spectrograms are averaged across patients separately for contralateral (upper panel) and ipsilateral (lower panel) sensorimotor cortex for each EEG session (Pre, Post1, Post2). The right hand panel displays overlaid beta power traces for the three sessions. The black rectangles indicate the time windows of interest of peak changes in beta activity (MRBD, PMBR). Please note that PMBR occurred at lower beta frequencies (10–25 Hz) compared to MRBD, in line with known age-related reduction beta peak frequency (Rossiter et al., 2014b). These time-frequency windows were identical for healthy age-matched controls (see (Espenhahn et al., 2019)), and tested for significant differences between groups and EEG sessions.

### 2.6 EEG recording

Scalp EEG (ANT Neuro, Asalab, The Netherlands) was continuously recorded at 2084Hz using 64 electrodes mounted on an elastic cap (waveguard EEG cap). The impedance was kept below ≤5kΩ and the EEG signal was referenced to Cz during recording. The timing of the visual cue (blue target) in the simple motor task was marked in the simultaneous EEG recording, with separate markers for each condition (flexion, extension). Surface electromyography (EMG) using bipolar electrodes in a belly-tendon montage placed on the wrist extensor (extensor carpi radialis longus) and flexor (flexor carpi radialis) muscles monitored movements of the affected hand.

### 2.7 Data analysis

#### 2.7.1 Motor learning

Motor performance on the continuous tracking task was parametrized by Root Mean Square Error (RMSE), an established measure implemented by other motor learning studies (Boyd and Winstein, 2006; Siengsukon and Boyd, 2009; Al-Sharman and Siengsukon, 2014; Espenhahn *et al*., 2019). RMSE captures the deviation of the wrist position at time *i* from the target position, and serves as a composite measure of temporal and spatial measurements of time lag and distance. RMSE was averaged across each block of training and retest sessions, with smaller RMSE values reflecting better motor performance. A linear regression model was fitted across the first and last 5 blocks of individual training and retest sessions to provide a performance estimate corrected for temporary effects such as fatigue or boredom (Adams, 1961) (as done previously by (Waters-Metenier *et al*., 2014; Espenhahn *et al*., 2019); see **Supplementary Figure 1**).

The analysis then concentrated on six time points in order to assess changes in motor performance across time: first block of training (T0), last block of training (T1), first block of retest1 (T2), last block of retest1 (T3), first block of retest2 (T4), and last block of retest2 (T5).

#### 2.7.2 Spectral power

Pre-processing and time-frequency analysis of EEG data during the performance of the simple motor task were performed using SPM12 (Wellcome Centre for Human Neuroimaging, http://fil.ion.ucl.ac.uk/spm) and additional scripts written in Matlab (version R2016a; The MathWorks Inc., Natick, MA, USA). The raw EEG signal was offline re-referenced to the average signal across all electrodes, bandpass filtered between 5–100Hz, additionally filtered with a 50Hz notch filter, and downsampled to 300Hz. Data were epoched from −1 to 9s relative to visual cue onset (0s). Poorly performed trials (e.g. movement initiated before cue signal) or those containing artefacts (e.g. eye blinks) were excluded. Artefact-free EEG time-series were decomposed into their time-frequency representations in the 5–45Hz range with frequency steps of 0.1Hz. A 7-cycle Morlet wavelet was used for the continuous wavelet transformation. Power was averaged across trials and rescaled in order to show changes relative to the corresponding pre-movement baseline period (−1–0s prior to cue onset), expressed as percentage of this baseline power.

Spectral power time-series were derived from a pre-selection of electrodes based on prior findings (Espenhahn *et al*., 2016) showing that the most prominent movement-related changes in beta activity for this simple motor task were observed in the following electrodes overlying the sensorimotor cortices contra- and ipsilateral to the trained wrist: ‘C4’ ‘CP4’ ‘CP2’ and ‘C3’ ‘CP3’ ‘CP1’ during movement-related beta desynchronization (MRBD); and ‘C2’ ‘C4’ ‘CP4’ and ‘C1’ ‘C3’ ‘CP3’ during post-movement beta rebound (PMBR). These bilateral electrodes were combined within hemispheres to derive resting beta power.

We chose specific time-frequency windows of interest based on peak changes in beta activity in time-frequency maps of the bilateral sensorimotor regions, which revealed clear movement-related beta-band (15–30Hz) activity in two distinct time windows of interest. This information was used to optimize the alignment of constant duration (1s) and width (15Hz) time-frequency windows to capture maximum MRBD (1–2s relative to cue onset), occurring between cue onset and movement termination, and PMBR (6–7s relative to cue onset), which emerges after movement cessation (**Figure 2**). These time-frequency windows were appropriate for patients as well as controls (see Figure 4 in (Espenhahn *et al*., 2019) for movement-related changes in spectral power in controls).

**Figure 3:**
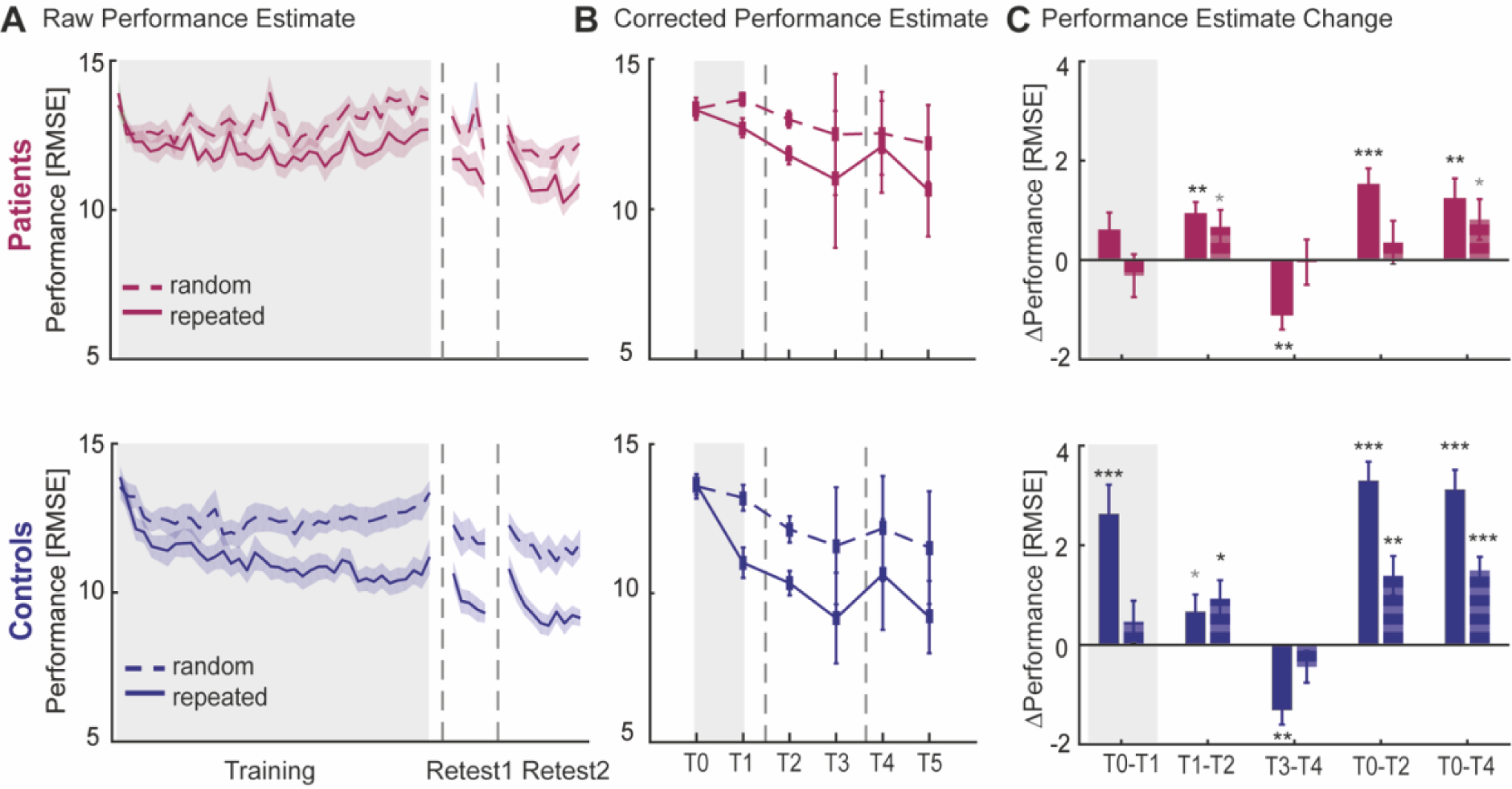
Motor skill learning of chronic stroke patients and age- and sex-matched healthy controls. **A**, Average motor performance (RMSE) for repeated and random sequences (solid and dashed lines respectively) across training (day 1), retest1 (day 1) and retest2 (day 2) sessions suggest reduced performance improvements of stroke patients (purple). Vertical dashed lines represent breaks between each session. **B**, Corrected performance estimates at the beginning and end of training (T0, T1) and retest (retest1: T2, T3; retest2: T4, T5) sessions. **C**, Performance differences (Δ) between time points, focusing on online learning (T0-T1) and offline learning across a shorter (retest1, T1-T2) or longer (retest2, T3-T4) time delay as well as overall performance changes from baseline (T0-T2; T0-T4). Solid bars represent Δ performance on the repeated sequence and striped bars on the random sequence. Positive and negative values, respectively, signify performance improvement and decrement. Shaded area (**A**) and error bars (**B, C**) indicate between-subject SEM. Statistical difference from zero:*p<0.05, **p<0.01, ***p<0.001, grey * p<0.1 (trend).

**Figure 4:**
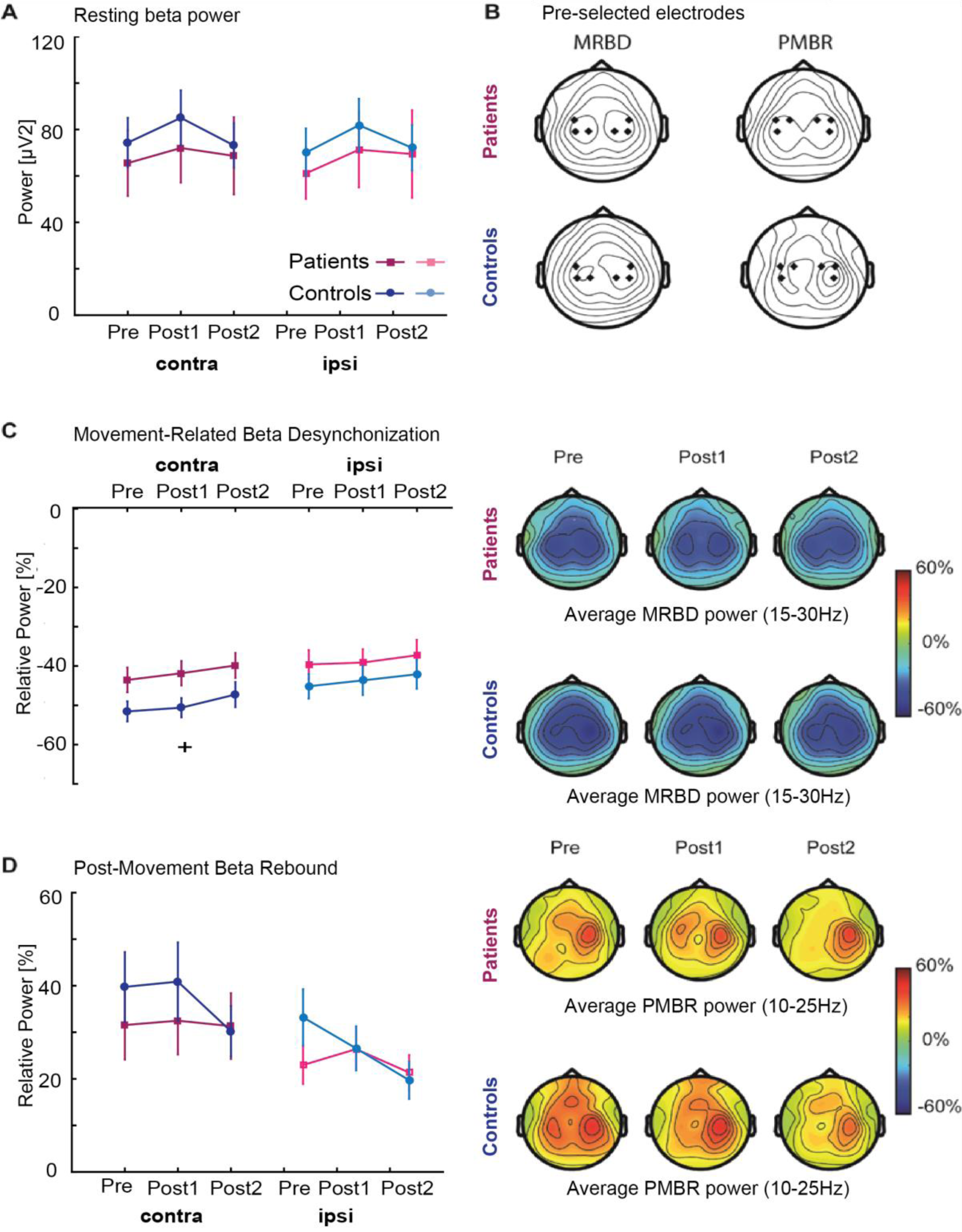
Alterations in beta power and corresponding topographic maps **A**, Average pre-movement (resting; −1–0s) beta power was comparable between patients (dark and light purple) and healthy controls (dark and light blue) for both sensorimotor cortices before (Pre), immediately after (Post1), and 24-hours after (Post2) training. **B**, Topographical plots of grand-averaged beta power showing the pre-selected electrodes (black diamonds) which were pooled as contralateral and ipsilateral regions of interest. **C-D**, Power in the movement (1–2s;MRBD) and post-movement time window (6-7s; PMBR) before (Pre), immediately after (Post1), and 24-hours after (Post2) training derived from contralateral and ipsilateral sensorimotor cortices of stroke patients (dark and light purple) and controls (dark and light blue) indicated no differential effect of stroke upon these beta dynamics. Error bars indicate between-subject SEM. Significant between-group differences are indicated with a ‘+’. Topographical distributions (right panels) of movement-related beta activity show differential contralateral and ipsilateral modulation patterns for MRBD and PMBR.

MRBD and PMBR were extracted from the respective 1s time windows and averaged for each EEG session (Pre, Post1, Post2) for the pre-selected electrodes over each hemisphere. The absolute pre-movement (resting) baseline beta (BB) power from −1 to 0s relative to cue onset was also obtained.

In total, 6 different beta parameter estimates were used for subsequent analyses: pre-movement baseline beta (absolute power), MRBD (relative power) and PMBR (relative power) from contra- and ipsilateral sensorimotor cortices, respectively.

### 2.8 Statistical analysis

Firstly, we examined effects of group, sequence type and time on motor performance parameters using a mixed-design ANOVA, with ‘group’ (2 levels: patients vs controls) as between-subject factor and ‘sequence type’ (2 levels: repeated vs random) and ‘time’ (5 levels: T0 vs T1 vs T2 vs T3 vs T4) as within-subject factors. Secondly, we examined effects of group, hemisphere and time on beta parameters using a mixed-design ANOVA, with ‘group’ (2 levels: patients vs controls) as between-subject factor and ‘hemisphere’ (2 levels: contralateral vs ipsilateral) and EEG ‘session’ (3 levels: Pre vs Post1 vs Post2) as within-subject factors. *Post-hoc* Bonferroni-adjusted *t*-tests were performed whenever main effects and interactions were found. Parametric tests were used as all variables were normally distributed.

Thirdly, in order to identify predictors of motor performance at T2 or T4 in our patient group, accounting for multicollinearity between measures, we used a multiple linear regression approach with stepwise selection (forward and backward algorithm; inclusion/exclusion probability levels: αEnter<0.05/ αExclude>0.1). We chose motor performance at T2 rather than T1 as it most likely reflects fairly stable learning effects unaffected by training-induced temporary effects such as fatigue or boredom (Rickard *et al*., 2008; Brawn *et al*., 2010), while performance at T4 indexes retention of the acquired motor skill overnight, reflecting motor memory consolidation (Robertson *et al*., 2005; Walker, 2005; Hotermans *et al*., 2006). A combination of spectral power measures, including (a) baseline beta power, (b) MRBD, and (c) PMBR from both sensorimotor cortices, as well as motor performance measures during the training session, i.e. (d) at T0 and (e) at T1, were used to explain performance at T2, while motor performance measures during retest1, i.e. (f) at T2 and (g) T3, were further included to explain performance at T4. In addition, demographic information such as age, motor function, cognitive function and sleep characteristics were equally included. See **Supplementary Table 2** for a full list of predictor variables included. All variables were z-scored before analysis to produce regression coefficients (β) of comparable magnitude and a leave-one-out cross-validation (LOOCV) approach was employed (Picard and Cook, 1984; Arlot and Celisse, 2010) to avoid overfitting and evaluate the predictive strength of each regression model. This cross-validation method is an established procedure for assessing generalization of results to an independent data set, particularly with smaller sample sizes (Huang *et al*., 2011; Kang *et al*., 2014). The strength of the prediction model was quantified in terms of the correlation coefficient between actual and predicted motor performance. A permutation-test (100 iterations) was used to assess whether the difference between the actual and predicted performance was greater than would be expected by chance (*p*-value below 0.05). All data in the main text and tables are presented as mean ±SD unless stated otherwise. Statistical analyses were performed using SPSS (version 22; IBM) and custom-written Matlab routines.

### 2.9 Data availability

The data supporting the findings in this study are available upon reasonable request from the corresponding author, S.E.

## 3 Results

All subjects were able to undergo training on the continuous tracking task and perform the simple motor task during EEG recording. The patient group studied here was well-recovered given their low level of impairment (**Supplementary Table 1**) and comparable motor and cognitive function to age-matched healthy controls (**Table 1**). Stroke patients only significantly differed from controls with regard to their sleep quantity for which they on average reported one hour of sleep more.

**Table 1:** Group characteristics of stroke patients and healthy controls.

|  | Patients | Controls | Between-group<br>difference |
| --- | --- | --- | --- |
| Handedness (Edinburgh) | 87±24 | 85±21 | $t_{(34)}=-0.21, p=0.833$ |
| Grip Strength [lb] | 66±26.04 | 63±21.03 | $t_{(34)}=0.41, p=0.682$ |
| NHPT [pegs/s] | 0.57±0.13 | 0.60±0.07 | $t_{(34)}=-0.93, p=0.362$ |
| SART (Error score, 0-225) | 13±8.97 | 13±10.73 | $t_{(34)}=0.13, p=0.897$ |
| SART (RT in ms) | 456±114.3 | 451±142.9 | $t_{(34)}=0.108, p=0.915$ |
| Sleep Quantity [hours]# | 7±1.02 | 6±0.94 | **$U=93.5, p=0.033$** |
| Sleep Quality (1-8)# | 4.7±1.57 | 5.2±0.87 | $U=141.0, p=0.560$ |

*Between-group comparisons only revealed a significant difference in sleep quantity. Independent-samples $t$ -tests were used to test for between-group differences. For discrete data (#), Mann-Whitney $U$ -tests were applied. Handedness was assessed using the Edinburgh Handedness Inventory (Oldfield, 1971). Motor functions are affected hand/non-dominant hand only and sleep measures are averaged across both days (both sleep measures were not significantly different between day 1 and day 2, $p>0.1$ ). Significant effects are indicated in bold. NHPT: Nine Hole Peg Test; SART: Sustained Attention to Response Test.*

### 3.1 Is motor skill learning altered after stroke?

Motor performance for both chronic stroke patients and healthy controls at training and retest sessions is shown in **Figure 3**A. We were able to directly compare performance on the motor learning task between groups because no systematic differences in baseline (block 1) performance between patients and controls [F_(1,34)_=0.42, *p*=0.523] or repeated and random sequences [F_(1,34)_=0.002, *p*=0.969] nor an interaction effect [F_(1,34)_=0.051, *p*=0.823] (**Figure 3**B) were present.

The mixed-design ANOVA on motor performance revealed a significant main effect of ‘time’ [*F*_(4,136)_=32.33, *p*<0.001, effect size *ŋ*_*p2*_=0.487], ‘sequence type’ [*F*_(1,34)_=55.216, *p*<0.001, effect size *ŋ*_*p2*_=0.619] and ‘group’ [*F*_(1,34)_=4.80, *p*=0.035, effect size *ŋ*_*p2*_=0.124]. In addition, we found significant interactions between ‘time x group’ [*F*_(4,136)_=4.25, *p*=0.006, effect size *ŋ*_*p2*_=0.111], ‘time x sequence type’ [*F*_(4,136)_=10.98, *p*<0.001, effect size *ŋ*_*p2*_=0.244], and ‘sequence type x group’ [*F*_(1,34)_=5.58, *p*=0.024, effect size *ŋ*_*p2*_=0.141], but no significant 3-way interaction was found. *Post hoc* analyses were performed separately and described in the following sections.

#### 3.1.1 Performance changes over the course of training

In contrast to the healthy age-matched controls, stroke patients did not show significant immediate improvements in motor performance with training (T0 vs T1) [*F*-statistics and *p*-values of ANOVAs are summarized in **Supplementary Table 3**], neither for the repeated [*t*_*(15)*_=1.62, *p*=0.127] nor random sequence [*t*_*(15)*_=-0.73, *p*=0.476]. Closer inspection of the tracking performance in **Figure 3**A shows a decline in performance towards the end of the training phase for the stroke patients, suggesting that temporary effects such as fatigue or boredom might have depressed performance towards the end of training.

#### 3.1.2 Performance changes after training

During the short time period between the end of the initial training and retest1 session (T1 vs T2), patients’ motor performance significantly improved by 7%, without further training, but only for the repeated sequence [*t*_*(15)*_=3.72, *p*=0.002, effect size *ŋ*_*p2*_=0.480]. This indicates a boost in performance early after the initial training (45-60 min) that did not significantly differ from healthy controls [*t*_(34)_=0.56, *p*=0.582] (**Figure 3**C).

In line, patients’ overall performance significantly improved from T0 to T2 for the repeated sequence only (11% improvement) [*t*_*(15*)_=4.53, *p*<0.001]. Together, this suggests that patients actually learned, but that the learning effects were masked at the end of training (T1), most likely due to temporary effects of fatigue. However, learning-related improvements were ∼50% smaller compared to the healthy control group [*t*_*(34)*_=-3.55, *p*=0.001].

Lastly, changes in motor performance, without practice, at 24 hours (retest2) after initial training were assessed. Overnight (T3 vs T4), stroke patients suffered a significant 10% performance decrease (i.e. forgetting) specific to the repeated sequence [*t*_*(15)*_=-3.51, *p*=0.003], which was similar to the 12% performance decrement observed in healthy controls [*t*_*(34)*_=0.01, *p*=0.992] (**Figure 3**C). Overall, stroke patients demonstrated significantly improved performance on the repeated sequence at T4 compared to T0 (9% improvement) [*t*_*(15)*_=2.91, *p*=0.011], but nevertheless their overall sequence-specific performance improvements were significantly smaller compared to healthy controls [*t*_*(34)*_=-3.67, *p*=0.001].

In summary, whilst capacity to learn a motor skill is preserved in our stroke patients, the rate of learning is diminished in comparison to healthy controls.

### 3.2 Do beta oscillations change with training after stroke?

Average spectral changes in contralateral and ipsilateral sensorimotor cortices in response to wrist movement are shown in **Figure 2** before (Pre) and at the two time points (Post1, Post2 – **Figure 1**A) after initial training. General features of the spectral changes in beta activity induced by the simple motor task have been detailed in a previous study (Espenhahn *et al*., 2016) and replicated in the elderly (Espenhahn *et al*., 2019).

#### 3.2.1 Resting beta power

Absolute pre-movement (resting) beta power in either contralateral or ipsilateral sensorimotor cortices was not different between stroke patients and age-matched healthy controls as evidenced by a lack of significant Group and Hemisphere effects (**Figure 4**A, *F*-statistics and *p*-values of all ANOVAs are summarized in **Supplementary Table 4**), consistent with previous observations (Rossiter *et al*., 2014*a*). However, absolute pre-movement (resting) beta power did change significantly across sessions. *Post-hoc* analyses revealed a significant but transient increase in beta power immediately after training (Post1) in both contra- [*F*_*(2,19)*_=5.93, *p*=0.006, *effect size ŋ*_*p2*_*=*0.238] and ipsilateral cortices [*F*_*(2,19)*_=7.67, *p*=0.002, *effect size ŋ*_*p2*_*=*0.287] in controls, which returned back to pre-training levels on day 2. This effect was not seen in stroke patients [*F*_*(2,30)*_=1.45, *p*=0.250].

#### 3.2.2 Movement-related beta power changes

MRBD and PMBR in both sensorimotor cortices and topographic maps are shown in **Figure 4**C-D. Interestingly, although the magnitude of MRBD was on average ∼10% smaller in stroke patients compared to controls, overall no significant group differences for either the contra-nor ipsilateral sensorimotor cortex were found (except for the contralateral side at time point post1) (**Figure 4**C). Similarly, estimates of PMBR were comparable between stroke patients and age-matched healthy controls (**Figure 4**D). In addition, both MRBD and PMBR significantly changed across sessions. *Post-hoc* analyses revealed a significant reduction across sessions in contralateral sensorimotor cortex for MRBD [*F*_*(2,19)*_=4.38, *p*=0.019, *ŋ*_*p2*_=0.187] and ipsilateral sensorimotor cortex for PMBR [*F*_*(2,19)*_=5.85, *p*=0.006, *ŋ*_*p2*_=0.235] in the healthy controls. Crucially, this training-related modulation of MRBD and PMBR was not evident in the stroke patients.

In summary, just as with motor performance, there were no significant differences in the properties of beta oscillations prior to training between stroke patients and healthy controls. However, less change in estimates of beta activity was observed across training (day 1 and 2) in our patients in comparison to controls.

### 3.3 Do beta oscillations predict post-training performance in stroke patients?

In order to determine whether there were significant predictors of skill learning at T2 or skill retention at T4 in our patient group, we employed a stepwise linear regression approach within a LOOCV.

Firstly, none of the factors listed in **Supplementary Table 2** significantly predicted motor performance shortly after training (T2). However, attempts to predict motor performance at T4 yielded a model with five significant predictive factors that accounted for 82% of the variance in motor performance 24 hours after initial training (T4) (**Figure 5**A). As expected, earlier motor behaviours (at T2 and T3) were the best predictors [T2: *β*=0.41, *t*_(15)_=6.43, *p*<0.001; T3: *β*=0.62, *t*_(15)_=9.67, *p*<0.001]. However, lower contralateral (ipsilesional) PMBR immediately after training (Post1) was associated with better future motor performance [*β*=0.21, *t*_(15)_=4.79, *p*<0.001]. In addition, dominance of the affected hand [*β*=0.13, *t*_(15)_=3.07, *p*=0.01] and sleep [*β*=-0.16, *t*_(37)_=-3.96, *p*<0.01] were additional explanatory factors. Similarly, *post-hoc* pairwise correlations revealed a non-significant correlation between post-training contralateral (ipsilesional) sensorimotor cortex PMBR and performance at T4 [r=0.10, *p*=0.711], which becomes significant after regressing out prior performance, hand dominance, and sleep as confounding covariates [squared semi-partial correlation: *r*^*2*^=0.62, *p*<0.001].

**Figure 5:**
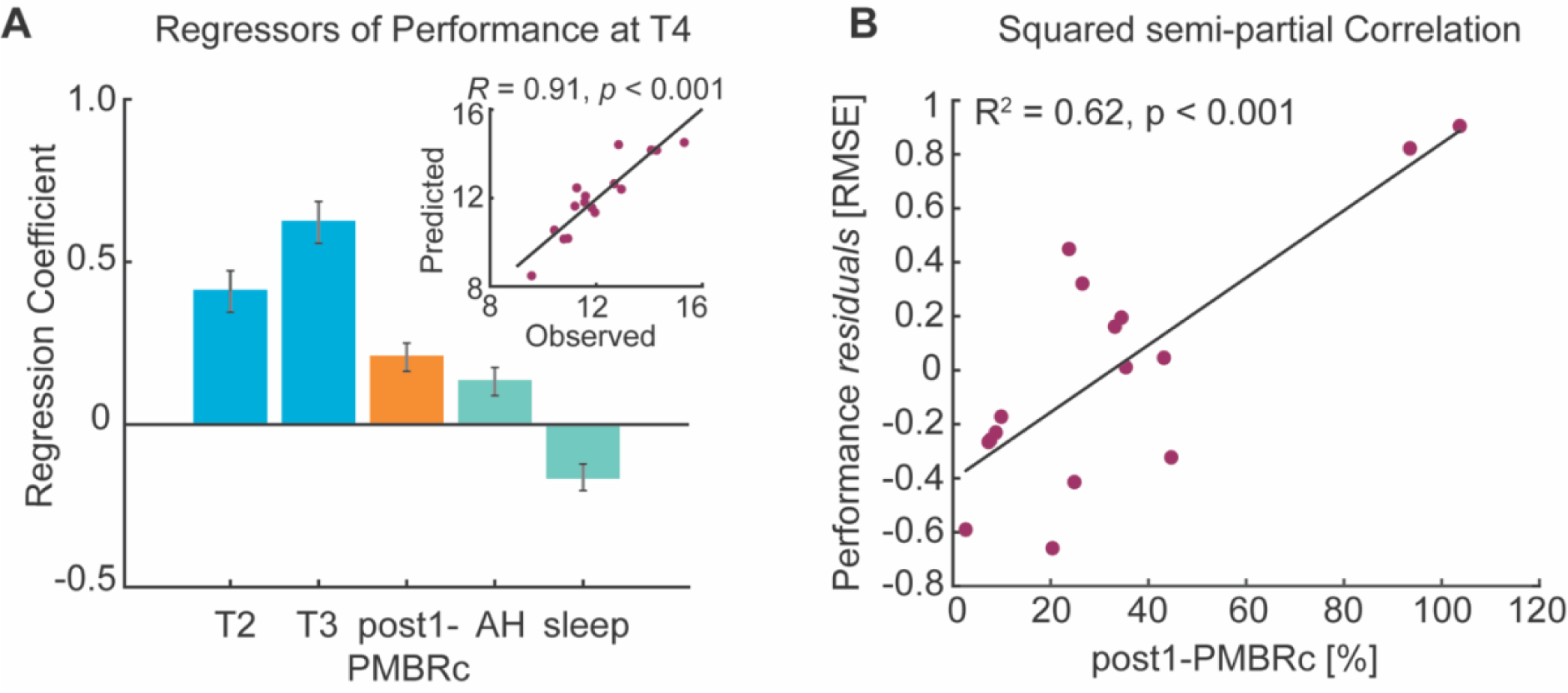
Prediction of motor performance at T4. Regression analysis provided statistically significant performance prediction (A) as quantified by the correlation between actual and predicted motor performance in stroke patients (inlet figure), with significance determined by permutation-testing. The model consisted of 5 significant predictors accounting for 82% of variance in performance 24 hours after training (T4). Patients’ performance during training, post-training movement-related beta activity, affected hand and sleep quantity were related to performance at T4. Z-scored regression coefficients (β) quantify the influence of each significant predictor upon performance level at T4. Error bars represent SEM. **B**, Importantly, post-hoc squared semi-partial correlation confirmed that movement-related beta activity immediately after training was positively related to performance at T4, indicating that smaller magnitude of contralateral (ipsilesional) PMBR is associated with better future performance.

## 4 Discussion

In this study, we were able to confirm that the capacity for motor skill learning is preserved in chronic stroke patients, but the rate of learning was diminished compared to healthy controls even when the task is of equal difficulty for everyone. Furthermore, we were able to show that one aspect of cortical oscillatory behaviour in stroke patients, specifically immediate post-training PMBR from contralateral (ipsilesional) sensorimotor cortex, contributed significantly to predicting motor performance 24 hours after training.

Making the comparison between stroke patients and healthy control subjects is fraught with difficulty because of differences in pre-training performance between the two groups. In this study, we avoided these performance confounds by individually determining the velocity with which the target moved (in contrast to studies that use a fixed speed), thus ensuring that task difficulty was equal across groups and left enough room for improvement in performance. Our patients therefore had no discernible differences in motor performance to the age-matched healthy controls at the beginning of training. Consistent with other studies (e.g. (Platz *et al*., 1994; Winstein *et al*., 1999; Boyd and Winstein, 2001, 2004, 2006; Pohl *et al*., 2006; Vidoni and Boyd, 2009; Hardwick *et al*., 2017)), we found that stroke patients were able to improve their motor performance with training, suggesting preserved motor learning ability after stroke. Despite abnormal patterns of brain activity that occur after stroke (Chollet *et al*., 1991; Weiller *et al*., 1993; Marshall *et al*., 2000; Johansen-Berg *et al*., 2002; Ward *et al*., 2003), preserved ability to learn in stroke patients may likely be due to the distributed nature of the neural network supporting motor learning (Karni *et al*., 1995; Sanes and Donoghue, 2000; Doyon and Ungerleider, 2002). However, we found that the overall level of performance achieved by stroke patients with short-term training (T0 to T2 and T0 to T4) was significantly reduced compared to age-matched healthy controls. Although it is not possible to say whether prolonged training (i.e. weeks) by our stroke patients would have resulted in equivalent levels of performance to healthy controls or whether patients reach a performance plateau that remains categorically different to healthy adults, our results show that some aspect of learning was affected.

In this study, we have measured cortical beta oscillations as biomarkers of the potential for learning through plasticity mechanisms. Despite evidence for aberrant beta activity after stroke (Rossiter *et al*., 2014*a*; Shiner *et al*., 2015), we rather unexpectedly did not find significant stroke-related alterations in beta oscillations before training started. Given that effective recovery of motor function is associated with a normalization of brain activity back towards a pattern seen in healthy controls (Johansen-Berg *et al*., 2002; Ward *et al*., 2003), it appears likely that the lack of post-stoke alteration in beta dynamics is due to restitution of nearly ‘normal’ beta activity in our well-recovered patient cohort. However, we did see differences in beta oscillations between the two groups as motor training progressed. While healthy controls demonstrated a transient post-training increase in pre-movement (resting) beta activity and reductions in both contralateral MRBD and ipsilateral PMBR with training, stroke patients did not show comparable patterns, suggesting less flexible modulation of cortical beta power accompanying learning in stroke patients. The transient training-related modulation of beta power might be related to a increase of cortical inhibition that is akin to temporary suppression of cortical plasticity with motor learning (Rioult-Pedotti *et al*., 1998, 2000, 2007; Ziemann *et al*., 2004; Stefan *et al*., 2006; Rosenkranz *et al*., 2007; Cantarero *et al*., 2013). We might speculate that this physiological response is necessary for practice-dependent plasticity processes to occur, and if absent or reduced as observed in the stroke patients, corresponds to reduced motor learning ability.

To date, several studies have investigated the relationship between properties of cortical beta oscillations and post-stroke motor impairment (Hall et al., 2010b; Laaksonen et al., 2012; Rossiter et al., 2014a; Shiner et al., 2015; Thibaut et al., 2017), but to the best of our knowledge, no study has explored whether cortical beta oscillations are associated with motor learning capacity after stroke. By employing a regression approach with LOOCV, we were able to show that movement-related beta dynamics were associated with future motor performance in chronic stroke patients. Specifically, post-training contralateral (ipsilesional) PMBR contributed significantly to a model that predicted motor performance levels 24 hours after training. More specifically, patients who exhibited lower PMBR after training performed better on the repeated sequence 24 hours after training. Given the link between beta oscillations and cortical GABA tone (Hall et al., 2011, 2010a; Jensen et al., 2005; Muthukumaraswamy et al., 2013; Roopun et al., 2006; Yamawaki et al., 2008), smaller post-training PMBR likely reflects lower GABAergic inhibition (Laaksonen, 2012), and therefore higher potential for training dependent plasticity. This general interpretation is in line with magnetic resonance spectroscopy (MRS) and positron emission tomography (PET) studies reporting decreases in GABA levels being associated with better motor recovery after stroke (Blicher et al., 2015; Kim et al., 2014). In line with our previous work (Espenhahn *et al*., 2019), this finding generally supports the idea that neurophysiological measures can detect individual differences in a ‘brain state’ that influence the effects of behavioural training, and might be used in future modelling approaches to help predict response to treatment (Reinkensmeyer *et al*., 2016)).

Whilst the focus on well-recovered patients could be seen as a limitation of our study, we argue that the strength of this approach lies in the investigation of motor learning independent of potentially obscuring influences of motor impairments. Further, it clearly showed that well-recovered patients with ‘normal’ motor control remain different to healthy adults in terms of their ability to learn, most likely due to lesion-induced structural and functional changes in the neural networks supporting motor learning. Nevertheless, given the notion of increased potential for plasticity and heightened responsiveness to motor training during the early post-stroke phase (Cramer, 2008; Krakauer et al., 2012; Murphy and Corbett, 2009; Ward, 2017; Zeiler and Krakauer, 2013), further work is required to determine whether beta oscillations in acute stroke patients similarly relate to motor learning ability.

In conclusion, the current results extend previous findings on the contribution of accessible beta oscillatory measures in explaining how motor skills are acquired on an individual level, beyond information provided by behavioural scores. While cortical oscillations may be only one of several factors important for motor learning, they may have value as markers of cortical function and plasticity after stroke and may offer novel targets for therapeutic interventions.

## Acknowledgements

The authors are grateful to Archy de Berker for coding support and Joshua Hadwen for technical testing and assistance in EEG cap preparation.

